# Medicine carbon footprinting across National Health Service trusts in England, 2021–2024: a longitudinal study using Freedom of Information requests

**DOI:** 10.64898/2026.10.09.26365120

**Authors:** Haroon Taylor, Nazneen Rahman

## Abstract

**Introduction:** Medicines account for 20% of the National Health Service (NHS) carbon footprint, but it is unclear how consistently they are measured by individual NHS trusts. This study assessed medicine carbon footprint (MCF) practices across NHS trusts in England and whether these changed between 2021 and 2024.

**Methods:** Freedom of Information requests were sent to NHS trusts about MCF practices in 2021 and 2024. Responses were analysed to determine the proportion of trusts calculating MCFs, the medicines included and the emission factors used for spend-based calculations. Reported spend-based factors were compared with the median factor for each year to assess the effect of factor choice on MCFs.

**Results:** Of 196 trusts included in analysis, 178 (91%) responded for 2021 and 182 (93%) for 2024. Among responding trusts, 30 of 178 calculated MCFs in 2021 compared with 46 of 182 in 2024. Spend-based medicine footprinting increased from 19 trusts in 2021 to 31 in 2024. Ten different factors were used by 15 trusts in 2021 and 13 by 21 trusts in 2024. Reported factors ranged from 31% below to 258% above the median in 2024.

**Conclusions:** Medicine carbon footprinting remained uncommon, with only one-quarter of responding NHS trusts calculating a footprint in 2024. Among trusts using spend-based methods, wide variation in emission factors produced substantially different estimated MCFs. Clear national guidance on spend-based methodology could improve consistency in the short term, while greater use of standardised product-level approaches may better support medicine decarbonisation over time.

## Introduction

Healthcare systems both contribute to greenhouse gas (GHG) emissions and respond to the growing health impacts of climate change. The National Health Service (NHS) accounts for around 4-5% of England’s carbon footprint [1]. In 2020, the NHS became the first national health system to commit to net zero, aiming to achieve this goal for all emissions it can influence by 2045 [2].

Medicines are one of the largest sources of GHG emissions within the NHS. The 2020 assessment of the NHS carbon footprint estimated that medicines accounted for approximately 25% of total emissions [1]. The 2025 assessment updated this to 20%, with 3% from inhalers and anaesthetic gases and 17% from other medicines and prescribing, the single largest emissions category [3,4].

Emissions from this 17% were estimated using methods where expenditure on medicines is multiplied by an emission factor expressed as carbon dioxide emissions per pound spent. This method is referred to as spend-based modelling throughout this article, but is also known as environmentally extended input-output or top-down modelling. Spend-based estimates are influenced not only by changes in medicine use, but also by the emission factor selected and differences in expenditure caused by factors such as inflation or medicine pricing [5,6]. Therefore, medicine carbon footprints (MCFs) calculated using spend-based methods are a high-level proxy for the underlying emissions [7].

The emissions from the 3% attributed to inhalers and anaesthetics were estimated using product-specific activity-based methods (sometimes called bottom-up modelling) [3]. This allows estimates to be more closely aligned with decarbonisation actions. For example, desflurane is an anaesthetic gas with a higher carbon footprint than lower-carbon alternatives with equivalent clinical effectiveness. National reduction targets supported by product-specific estimates have led to a 98% reduction in desflurane use across the NHS between 2020 and 2025, reducing emissions by approximately 41 ktCO_2_e [4]. Similarly, estimates of the emissions of inhaler propellants showed that pressurised metered-dose inhalers have higher carbon footprints than alternatives such as dry-powder inhalers [8]. Increased use of clinically appropriate lower-carbon inhalers has contributed to a 340 ktCO_2_e (33%) reduction in emissions between 2020 and 2025 [4]. These examples demonstrate how product-specific quantification can support effective decarbonisation policies by directly measuring the emissions associated with changes in medicine use. Spend-based methods are not appropriate for this purpose because they estimate emissions from the amount spent purchasing medicines, rather than from the emissions associated with the products themselves.

Under the Health and Care Act 2022, NHS trusts have a duty to contribute towards UK net zero emissions targets [9]. To achieve this, every trust has a Green Plan which sets out the net zero initiatives they will implement in the next three years [10]. Medicines are highlighted as one of nine areas of focus, demonstrating their importance for achieving net zero [11]. Studies have assessed Green Plans and evaluated their effectiveness, but to our knowledge no study has focused specifically on MCFs [12–14].

The Freedom of Information (FOI) system allows any person or organisation to request information held by public bodies [15]. FOI requests are deployed as a research method to obtain information across NHS organisations, and have been used to investigate NHS trust sustainability practices [16,17]. FOI requests therefore provide an opportunity to ask all NHS trusts the same questions systematically at multiple defined time points to gain insight into MCF practices.

This study aimed to assess medicine carbon footprinting practices across NHS trusts in England, and how these changed between 2021 and 2024. We examined the proportion of trusts calculating MCFs, the medicines included in those calculations and, for trusts using spend-based methods, the emission factors applied. We also assessed the extent to which the choice of spend-based factor affected the resulting carbon footprint estimate.

## Methods

### Study design

We used the Freedom of Information (FOI) system to send requests to NHS trusts in England. We asked trusts about their medicine carbon footprint (MCF) practices using six questions with a mixture of multiple choice, numeric and free text responses available (Table 1). The first three questions asked about MCFs in general and focused on whether MCFs were calculated, what the total MCF was and which medicines were included in calculations. The last three questions focused on spend-based emission factors (we used the term “top-down” in the requests) and asked which factor was used, how often it was updated and why it was chosen.

**Table 1.** Freedom of Information request questions.

| No. | Question | Response format |
| --- | --- | --- |
| 1 | Do you calculate the carbon footprint of medicines? | **Single choice:** Yes; No |
| 2 | What was the carbon footprint of medicines for 2021/2024? | **Numerical value** in tonnes of CO<sub>2</sub>e, or other units if specified |
| 3 | How did you calculate the carbon footprint of medicines? | **Single choice:** Top-down; top-down + bottom-up; or other, with additional detail where applicable |
| 4 | For top-down calculations what factor did you use? | **Numerical value** in kg or tonnes of CO<sub>2</sub>e per £ spent, or other unit if specified |
| 5 | Do you update the factor used? | **Single choice:** updated yearly; same factor used every year; or updated less frequently than yearly |
| 6 | Why did you use this factor? | **Free-text response**, including when and by whom the factor was supplied |

### Data collection

We sent the request at two time points. The requests were identical, apart from the year specified. We sent the first request in October 2022 asking about MCF practices in 2021 and the second in October 2025 asking about 2024. Throughout this article, we refer to the request responses as the 2021 and 2024 responses, respectively.

We followed the FOI process for each individual trust using the details on their website. Trusts allow FOI requests to be made in three ways:

> Option A: By email to a specific FOI email address
>
> Option B: Using an online FOI form
>
> Option C: By post to the trust’s registered address

We used Option A where available and Option B otherwise.

We sent the 2021 request to 215 trusts and the 2024 request to 203 trusts. To enable comparison between the two time points, we reviewed organisational changes that occurred between 2021 and 2024. We retained trusts that continued to exist in 2024, including those that had changed name or absorbed another trust. We excluded trusts that had merged to form a new organisation or had been absorbed into another trust. This resulted in 196 trusts being included in the analysis.

### Data processing and analysis

We collated responses in Microsoft Excel, labelling trusts with their Organisation Data Service (ODS) codes for ease of reference. We standardised carbon footprints to tonnes of carbon dioxide equivalent (tCO2e) and spend-based emission factors to kilograms of carbon dioxide equivalent per pound spent (kgCO_2_e/£) if alternative units were provided. Additionally, we rounded spend-based emission factors to two decimal places.

We categorised responses to Question 3 into three non-mutually exclusive categories according to the medicines included in the carbon footprint calculation: inhalers, anaesthetic gases, and medicines calculated using spend-based methods. We assigned these categories using the selected response and the accompanying free-text information. All collated and standardised responses are provided in Supplementary file S1.

We calculated response rates for 2021 and 2024 separately. We then identified trusts that responded in both years to compare their responses between 2021 and 2024. We performed further analyses on the trusts that provided a spend-based emission factor in either year, assessing the number and distribution of emission factors used.

We analysed responses as provided and did not perform any independent verification. We performed the analysis in July 2026 using Python version 3.11 with pandas version 2.3 and NumPy version 1.26. Any responses received after this date were not considered.

### Sensitivity analysis

To assess the effect of factor choice on spend-based medicine carbon footprints (MCFs) we conducted a sensitivity analysis for 2021 and 2024. For each year, we calculated the median spend-based emission factor and the percentage difference between each trust’s reported factor and the median. Because spend-based MCF estimates are directly proportional to the emission factor used, this percentage difference represents the corresponding percentage change in the estimated MCF.

## Results

### Response rates

We assessed the NHS trust response rates to the FOI requests in 2021 and 2024. Of the 196 trusts included in the analysis, 178 (91%) responded in 2021 and 182 (93%) responded in 2024. 166 trusts responded in both years (85%), 12 responded in 2021 only, 16 responded in 2024 only and two did not respond in either year.

### Trusts calculating medicine carbon footprints

We assessed the number of NHS trusts that calculated medicine carbon footprints (MCFs) in 2021 and 2024 and how this changed over time. 30 of the 178 responding trusts (17%) in 2021 calculated MCFs, compared to 46 of 182 responding trusts (25%) in 2024. Among the 166 trusts who responded in both years, 112 (67%) did not calculate MCFs in either year, 10 (6%) calculated them in 2021 only, 25 (15%) in 2024 only and 19 (11%) in both years (Table 2).

**Table 2.** Change in medicine carbon footprinting between 2021 and 2024.

| Question 1: Do you calculate the carbon footprint of medicines? |  |  |
| --- | --- | --- |
|  | 2024: Yes | 2024: No |
| 2021: Yes | 19 | 10 |
| 2021: No | 25 | 112 |

*Includes trusts that responded to Question 1 in both years (n = 166).*

### Medicines included in carbon footprinting

We analysed which medicines were included in NHS trust carbon footprint calculations, using three categories: inhalers, anaesthetic gases and medicines assessed using spend-based methods. These categories were not mutually exclusive, so a single trust could calculate MCFs for zero, one, two or three categories. Of the 178 responding trusts in 2021, 17 (10%) calculated inhaler emissions, 16 (9%) anaesthetic gas emissions and 19 (11%) spend-based MCFs. In 2024, 19 of 182 trusts (10%) included inhalers, 23 (13%) anaesthetic gases and 31 (17%) spend-based MCFs (Figure 1). Five of 178 trusts (3%) in 2021 and 12 of 182 (7%) in 2024 calculated MCFs for all three categories. Overall, 148 of 178 trusts (83%) calculated MCFs for none of the categories in 2021, compared with 136 of 182 (75%) in 2024.

**Fig. 1.**
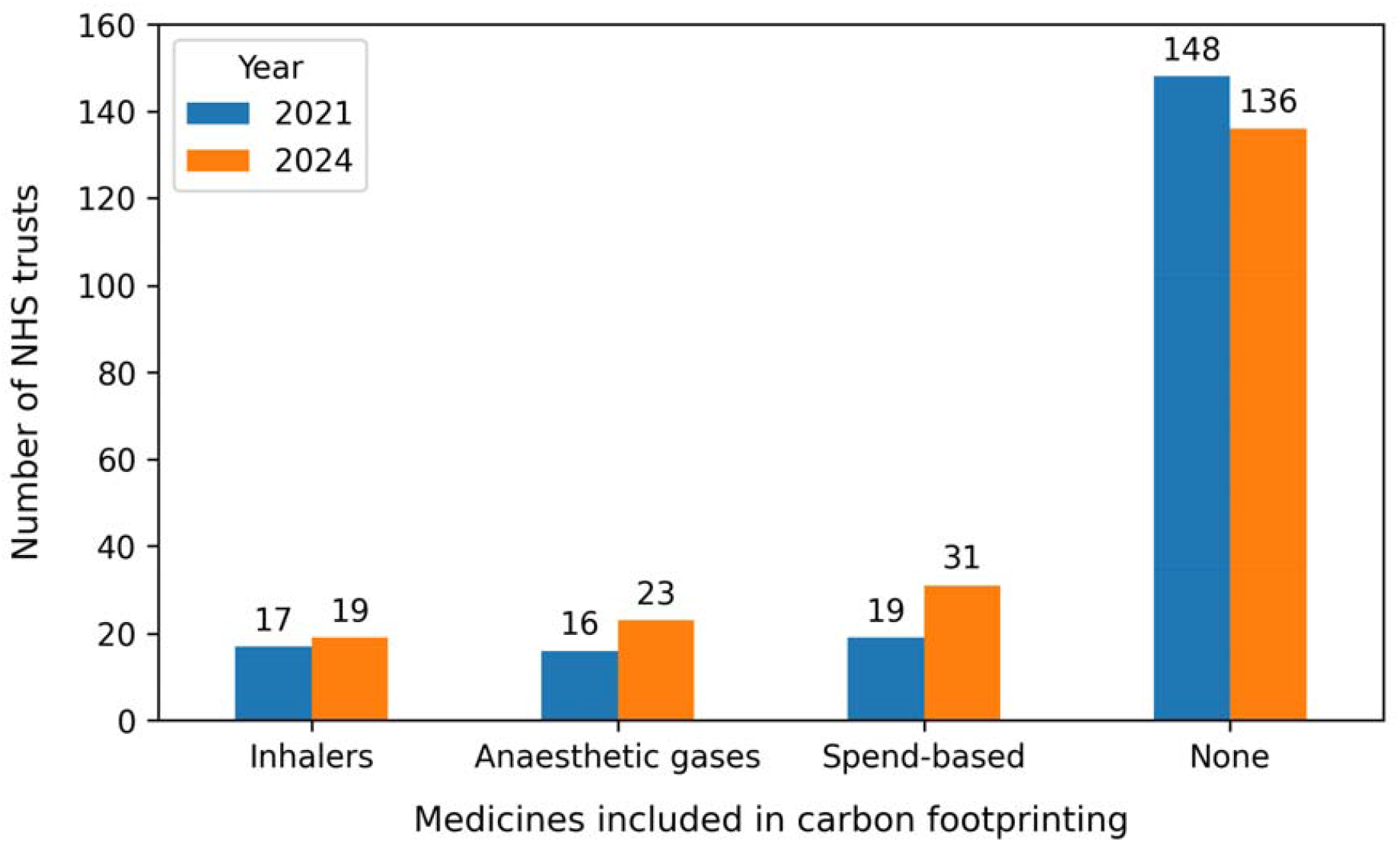
Number of NHS trusts calculating medicine carbon footprints for inhalers, anaesthetic gases and medicines assessed using spend-based methods in 2021 and 2024. Categories were not mutually exclusive; trusts could therefore be included in more than one category. “None” indicates trusts that did not calculate medicine carbon footprints for any of the three categories.

### Spend-based emission factors

We evaluated the spend-based emission factors reported by NHS trusts. In 2021, 15 of 19 trusts (79%) calculating spend-based MCFs provided the emission factor used. Ten different factors were reported, with 0.43 kgCO_2_e/£ the most common, used by four trusts. The median factor was 0.34 kgCO_2_e/£. In 2024, 21 of 31 trusts (68%) provided their emission factor. Thirteen different factors were reported, with 0.43, 0.51 and 0.62 kgCO_2_e/£ each used by three trusts. The median factor was 0.43 kgCO_2_e/£ (Figure 2).

**Fig. 2.**
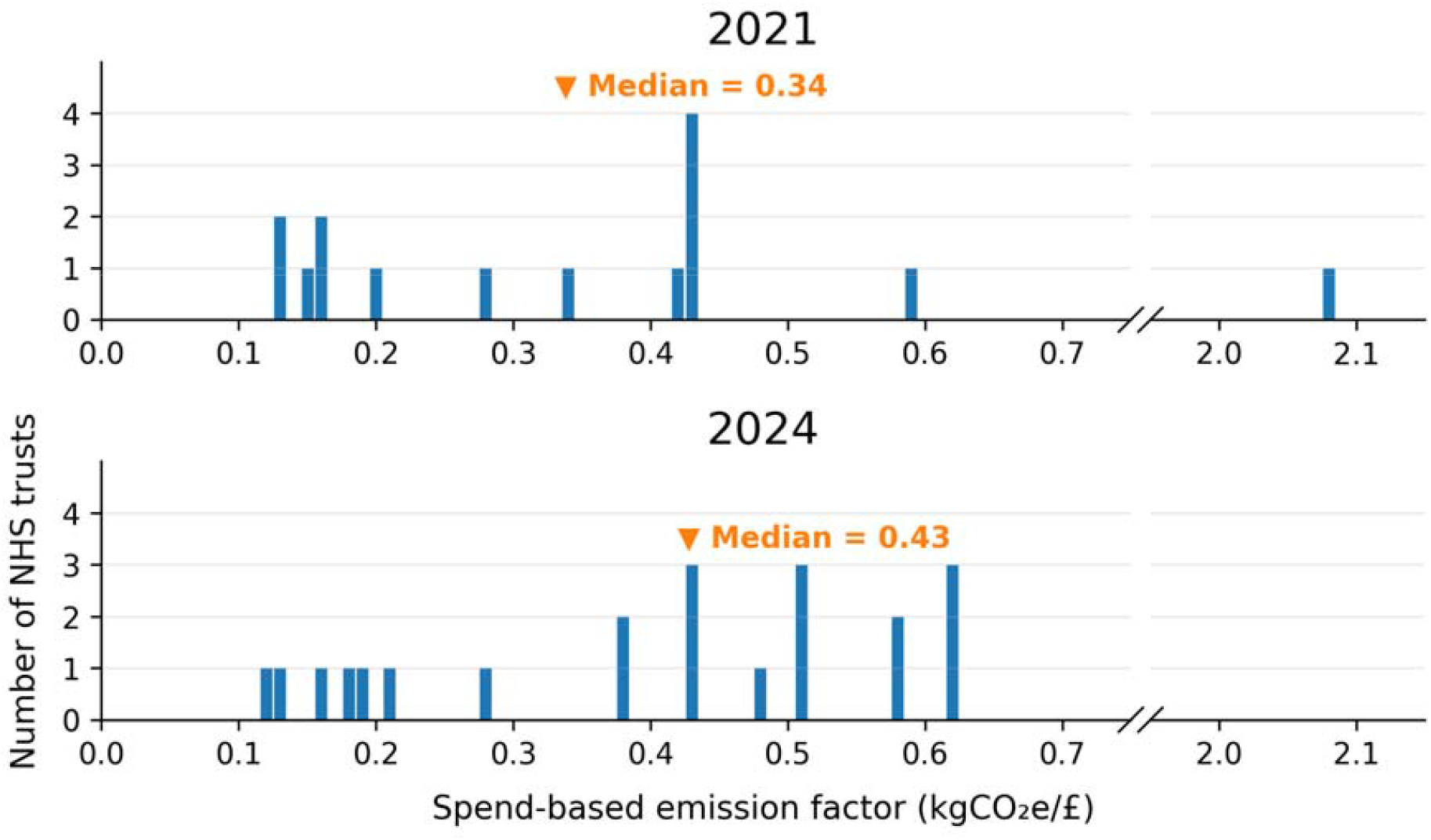
Distribution of spend-based emission factors reported by NHS trusts in 2021 and 2024. Orange markers indicate the median factor for each year. The x-axis is discontinuous between 0.7 and 2.0 kgCO_2_e/£ to accommodate an outlying value.

### Spend-based factor sensitivity analysis

To assess the effect of factor choice on spend-based MCF estimates, we compared each trust’s reported emission factor with the median factor for that year. In 2021, reported factors ranged from 84% below to 162% above the median; in 2024, they ranged from 31% below to 258% above the median (Figure 3). The highest and lowest factors differed by approximately 16-fold in 2021 and five-fold in 2024. Because spend-based MCF estimates are directly proportional to the emission factor used, these differences translate directly into equivalent percentage differences in the estimated MCF.

**Fig. 3.**
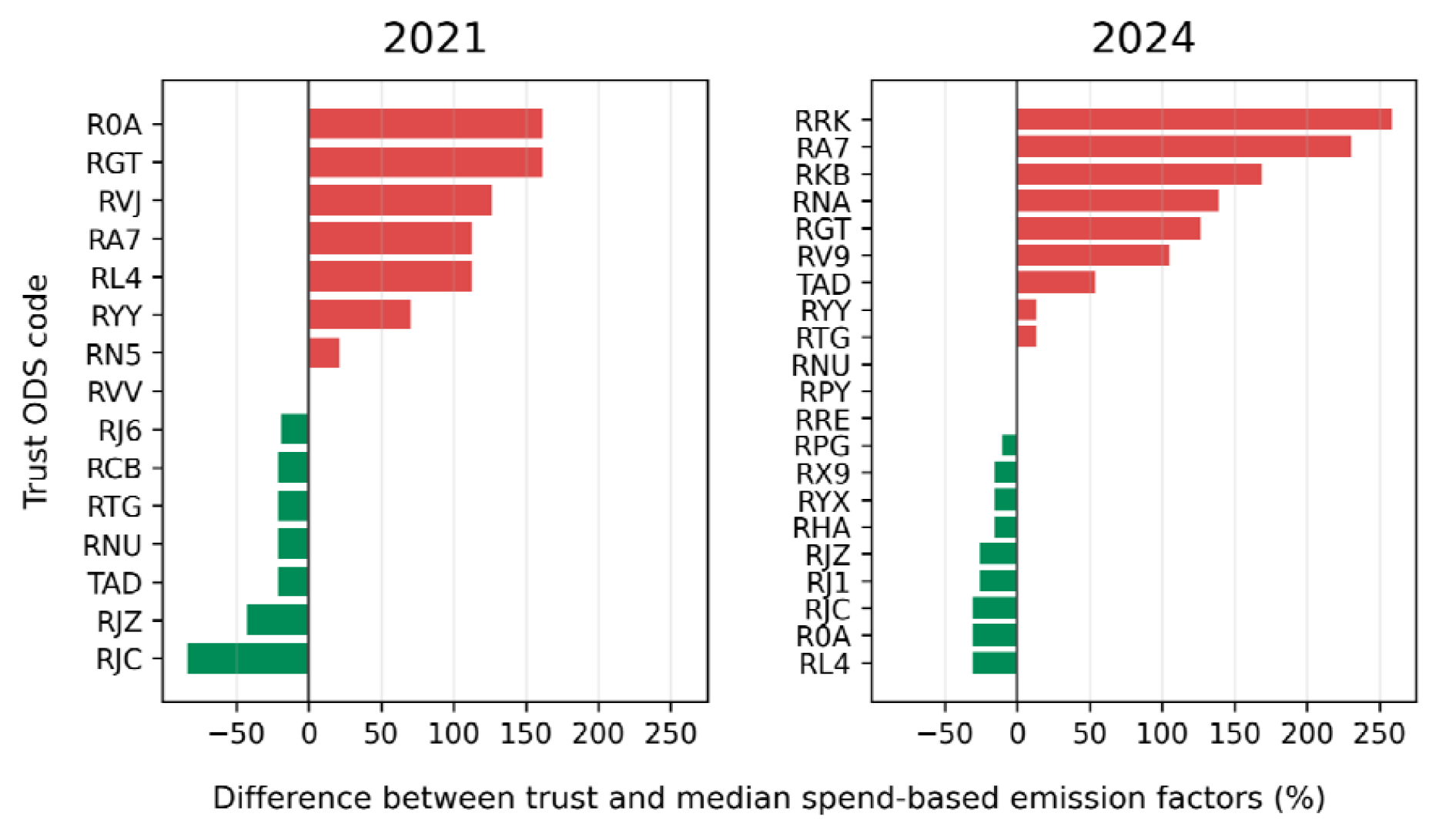
Percentage difference between the spend-based emission factors reported by NHS trusts and the median factors for 2021 and 2024. Trusts are denoted by Organisation Data Service (ODS) code.

## Discussion

In this study we provide a longitudinal assessment of how NHS trusts in England measure medicine carbon footprints (MCFs) in practice. We sent FOI requests to all available trusts regarding their MCFs in 2021 and 2024. We standardised responses and assessed organisational changes to ensure that results could be compared over time. We analysed the number of trusts that calculated MCFs, the categories of medicines that were footprinted and the spend-based emission factors that were used. We also assessed the effect of variation in spend-based factor choice using a sensitivity analysis. To our knowledge, this is the first study to focus specifically on MCF calculations by NHS trusts and one of the most comprehensive studies to ask trusts about sustainability practices.

We found that medicine carbon footprinting remained uncommon across NHS trusts. Three-quarters of responding trusts did not calculate MCFs in 2024. Although there was an increase of eight percentage points between 2021 and 2024 in the proportion of responding trusts calculating MCFs, this remained low given that medicines are the single largest contributor to NHS emissions and are identified as a focus area within NHS Green Plans [3,11]. These findings suggest that there remains a gap between national ambitions for reducing medicine-related emissions and routine measurement at trust level.

This finding is consistent with wider studies of sustainability planning across NHS trusts. An assessment of net-zero strategies across 214 NHS trusts highlighted the need for standardised measurement tools and key performance indicators to improve consistency and comparability [12]. Similarly, a study analysing 190 NHS Green Plans using the SMART framework found that fewer than half of strategies were “measurable” [13]. Our findings extend this evidence specifically to medicines, showing that MCF measurement is both limited in uptake and inconsistent in methodology.

Where medicine-specific targets and measurement approaches have been established nationally, substantial reductions have been achieved. For example, targeted action on desflurane and inhalers has been accompanied by large reductions in emissions from these sources [3]. This suggests that clearly defined medicine groups, consistent methods of measurement and measurable targets can support decarbonisation. Similar opportunities may exist for other medicine groups where reductions in use are already aligned with clinical priorities. For example, reducing unnecessary antibiotic use is an established antimicrobial stewardship goal, and antibiotic prescribing is already measured at trust level [18,19]. Medicine-specific carbon footprinting could therefore provide an additional way of identifying and evaluating such opportunities [20].

A further important finding was the substantial variation in spend-based emission factors used by NHS trusts. In 2024, the highest factor reported was approximately five times the lowest, and the sensitivity analysis showed that the estimated MCF could vary substantially depending on the factor selected. This limits the comparability of spend-based MCF estimates between trusts and over time.

The variation is likely to reflect the absence of a nationally agreed approach to selecting and updating spend-based emission factors.

Given the current use of spend-based methods, greater standardisation would improve the consistency of MCF measurement across NHS trusts. NHS England could provide guidance on the specific spend-based factor to use, how it should be applied and how frequently it should be updated. This would allow trusts choosing to calculate spend-based MCFs to generate more comparable estimates and would be consistent with the wider aim of improving measurement within Green Plans [21].

In the longer term, spend-based estimates have important limitations for measuring medicine decarbonisation. They depend on the data source, expenditure category or sector selected, and the year of the underlying data [22]. Medicine prices can also vary for reasons unrelated to environmental impact, meaning that changes in expenditure do not necessarily reflect changes in carbon emissions [7]. NHS England has acknowledged that reliance on spend-based emission factors limits its ability to measure progress in reducing emissions [4]. The NHS Net Zero Supplier Roadmap plans to introduce requirements from April 2028 for carbon footprint information for individual products supplied to the NHS [23], while standards such as PAS 2090 provide rules for pharmaceutical product carbon footprinting that go beyond spend-based estimates [24]. Together, these developments support a longer-term transition towards product- and activity-based approaches.

Increasing availability of standardised product-level MCF data may help enable this transition. For example, the MCF Product Dataset provides systematically calculated product-specific MCFs linked to the NHS dictionary of medicines and devices (dm+d) identifiers [25,26]. Such approaches could allow medicine emissions to be linked more directly to medicine use and therefore to specific decarbonisation interventions.

This study had several strengths. More than 90% of included NHS trusts responded in each year, providing broad national coverage of MCF practices. The same FOI questions were used at both time points, allowing us to assess changes over time. The use of FOI requests also allowed us to obtain information from trusts directly irrespective of their level of engagement with medicine carbon footprinting, rather than relying on participation in a voluntary sustainability survey.

This study also has limitations. We analysed FOI responses as provided by each trust and did not independently verify them against other sources. Responses may have been prepared by different staff at the two time points, potentially introducing variation in how questions were interpreted or information was retrieved. Additionally, the design of the questions introduced some uncertainty over the MCF timeframe. Some trusts reported data by calendar year and others by financial year, creating variation in the periods represented. The sample size for the sensitivity analysis was small, with only 15 trusts in 2021 and 21 trusts in 2024, reflecting the low levels of spend-based MCF calculation. Finally, in the absence of a nationally agreed standard factor, we used the median factor as a comparator rather than a validated reference value.

## Conclusion

In this study, we used Freedom of Information requests to assess medicine carbon footprinting practices across NHS trusts in England in 2021 and 2024. Medicine carbon footprinting remained uncommon, with three-quarters of responding trusts not calculating the carbon footprint of any medicines in 2024. There was also considerable variation in how medicine carbon footprints were calculated. Among trusts using spend-based methods, the emission factors used differed by approximately five-fold in 2024, limiting the comparability of estimates between organisations and over time.

Clear national guidance, including a recommended spend-based method and emission factor, could improve consistency in MCF measurement across NHS trusts. In the longer term, greater availability of standardised product-level medicine carbon footprint data linked to medicine quantity data could support identification and evaluation of medicine decarbonisation opportunities in line with NHS net zero goals.

## Data Availability

All data produced in the present study are available upon reasonable request to the authors

## List of abbreviations

CO_2_e: Carbon dioxide equivalent
dm+d: Dictionary of Medicines and Devices
FOI: Freedom of Information
GHG: Greenhouse gas
MCF: Medicine carbon footprint
NHS: National Health Service
ODS: Organisation Data Service

## Declarations

### Ethics approval and consent to participate

This study analysed information provided by NHS trusts in response to Freedom of Information requests. The data related to organisational practices and did not include identifiable personal data. Ethical approval and participant consent were therefore not required.

### Consent for publication

Not applicable.

### Availability of data and materials

All data supporting the findings of this study are available within the paper and its Supplementary Information.

### Competing interests

H.T. and N.R. are employees of and hold shares in YewMaker.

### Funding

This research did not receive any specific grant from funding agencies in the public, commercial, or not-for-profit sectors.

### Authors’ contributions

H.T. and N.R. undertook the study design, method development, data acquisition, data analysis and manuscript writing.

## Supplementary files

**Filename:** Supplementary file S1

**File format:** Excel (.xlsx)

**Title:** Freedom of Information responses on medicine carbon footprinting practices across NHS trusts.

**Description:** Processed trust-level data from the 2021 and 2024 Freedom of Information requests for the 196 NHS trusts included in the analysis. The dataset includes NHS trust names and Organisation Data Service codes, response status, and where available medicines included in carbon footprint calculations and reported spend-based emission factors.

## Notes

### Competing Interest Statement

The authors have declared no competing interest.

